## Supplementary material for "Enhancing surveillance of sexually transmitted infections in England with gender identity and behavioural data: the GUMCAD STI Surveillance System"

Supplementary Material – Table 1. Number of records submitted by sexual health services (SHS) that participated in the second pilot of the enhanced GUMCAD specification, England, 2015/16

| Clinic name | Number of records submitted |
| --- | --- |
| SHS 1 (East of England) | 963 |
| SHS 2 (South West) | 12,245 |
| SHS 3 (London) | 3,899 |
| SHS 4 (London) | 5,092 |
| SHS 5 (East of England) | 908 |
| Total | 23,107 |

Supplementary Material – Table 2. Feedback on the overall consistency, ease of completion, acceptability to patients, clarity of the technical guidance document and benefit of the proposed enhancement to GUMCAD, clinic staff from pilot sexual health services, 2016 (N=21)

| Variable | Agreed (%) | Not agreed (%) | Neutral (%) |
| --- | --- | --- | --- |
| Consistent with clinical practice | 43 | 19 | 38 |
| Ease of completion | 57 | 24 | 29 |
| Acceptable to patients | 71 | 10 | 19 |
| Clarity of the technical guidance document | 62 | 10 | 29 |
| Benefit of the proposed GUMCAD enhancement | 43 | 19 | 38 |

Source: Anonymous, self-administered online survey of clinicians who participated in the second pilot in 2015-16 (5 sexual health services participated in this pilot)

Supplementary Material – Table 3. Feedback on the ease of completion and usefulness of each of the four proposed behavioural domains of the enhanced GUMCAD specification, clinic staff from pilot sexual health services, 2016 (N=21)

|  | Response | Sexual behaviour (%) | Drug and alcohol use (%) | Previous STIs (%) | Partner notification (%) |
| --- | --- | --- | --- | --- | --- |
| Piloted variables were easy to complete |  |  |  |  |  |
|  | Yes | 81 | 48 | 48 | 10 |
|  | No | 10 | 33 | 33 | 0 |
|  | Not answered | 10 | 19 | 19 | 91 |
| Piloted variables were useful to collect |  |  |  |  |  |
|  | Yes | 81 | 57 | 62 | 10 |
|  | No | 5 | 19 | 14 | 0 |
|  | Not answered | 14 | 24 | 24 | 91 |

Source: Anonymous, self-administered online survey of clinicians who participated in the second pilot in 2015-16 (5 sexual health services participated in this pilot)

Supplementary Material –Figure 1. Survey instrument used to obtain feedback on the piloted enhanced GUMCAD specification after both the first and second pilots

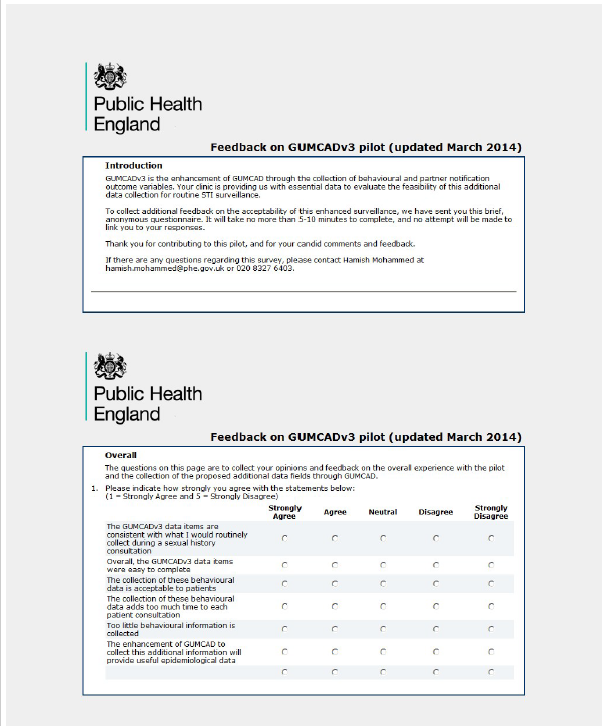

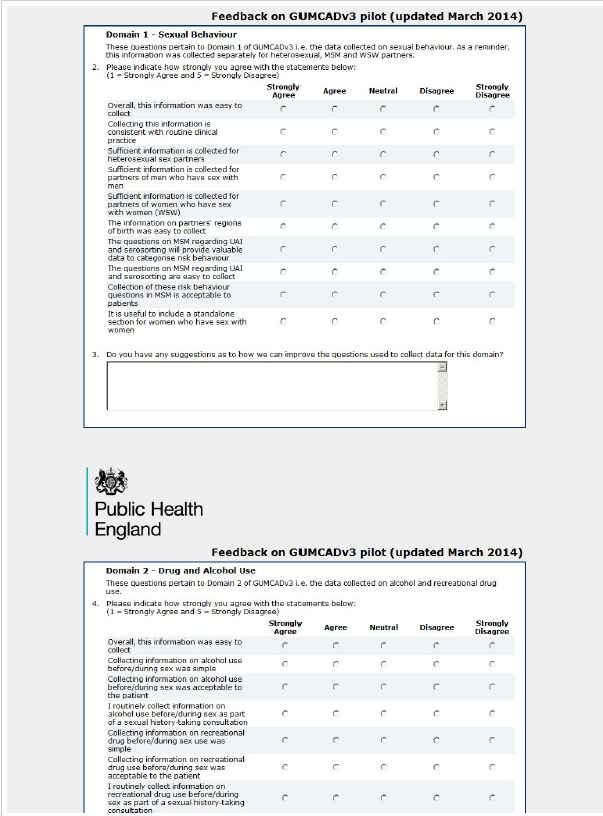

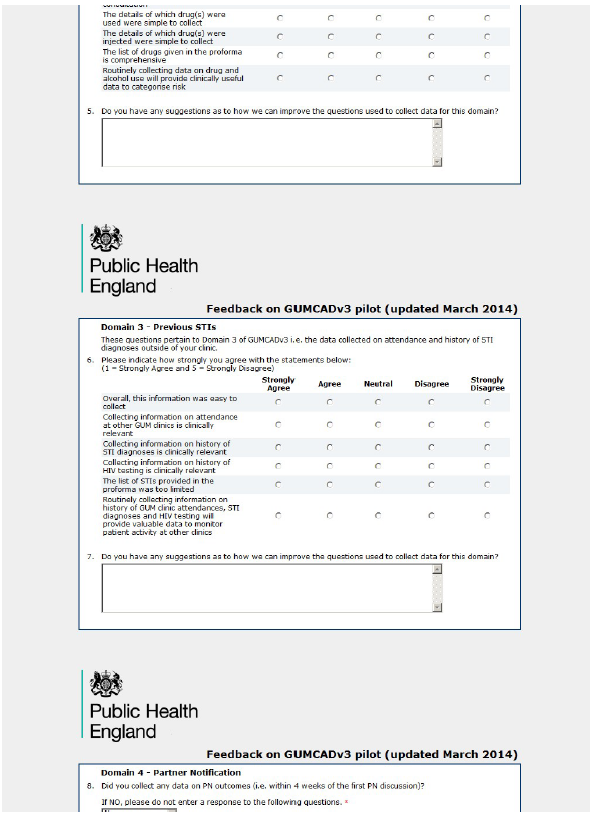

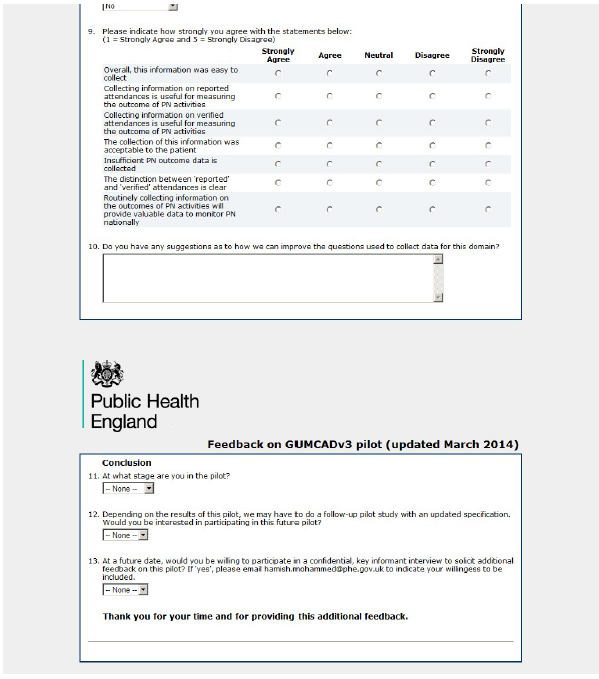

Notes: This survey was built in SelectSurvey. Public Health England is the predecessor to the UK Health Security Agency.

Supplementary Material – Figure 2. Proportion of patients attending 5 pilot sexual health services by gender and whether they had enhanced data submitted, pilot of enhanced GUMCAD specification, England, 2015/16

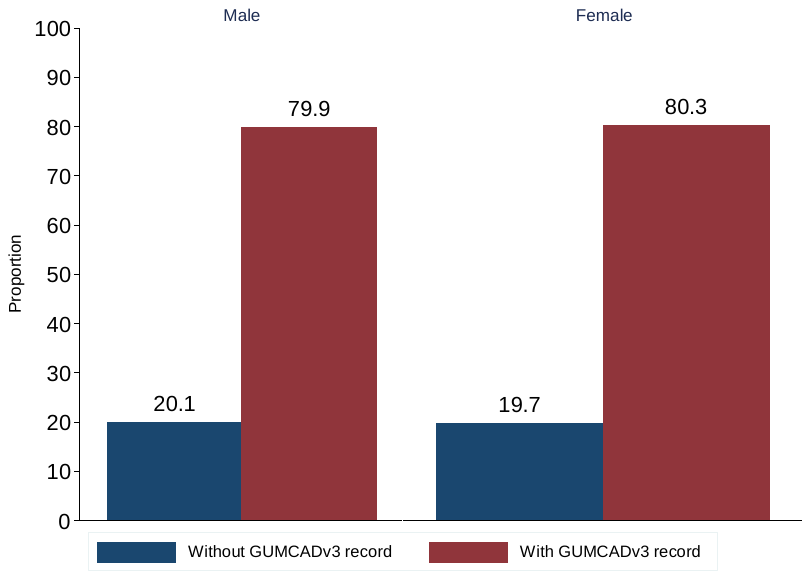

Key: GUMCADv3 – enhanced GUMCAD specification

The data collected in the pilot were based on a binary gender classification. The current version of GUMCAD collects data on gender identity (whether people are cisgender, transgender or gender-diverse).

Supplementary Material – Figure 3. Proportion of patients attending 5 pilot sexual health services by gender and age-group, and whether they had enhanced data submitted, pilot of enhanced GUMCAD specification, England, 2015/16
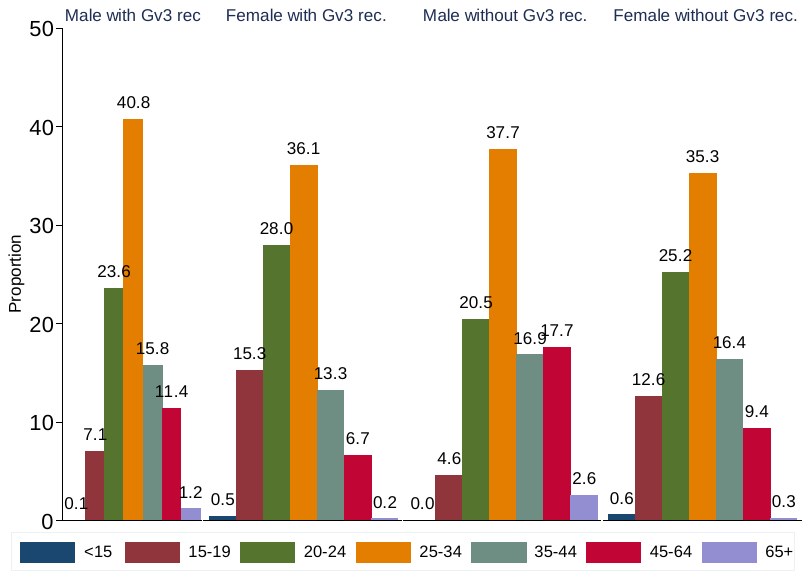

Key: Gv3 – enhanced GUMCAD specification

The data collected in the pilot were based on a binary gender classification. The current version of GUMCAD collects data on gender identity (whether people are cisgender, transgender or gender-diverse).

Supplementary Material – Figure 4. Proportion of patients attending 5 pilot sexual health services by gender and ethnicity, and whether they had enhanced data submitted, pilot of enhanced GUMCAD specification, England, 2015/16

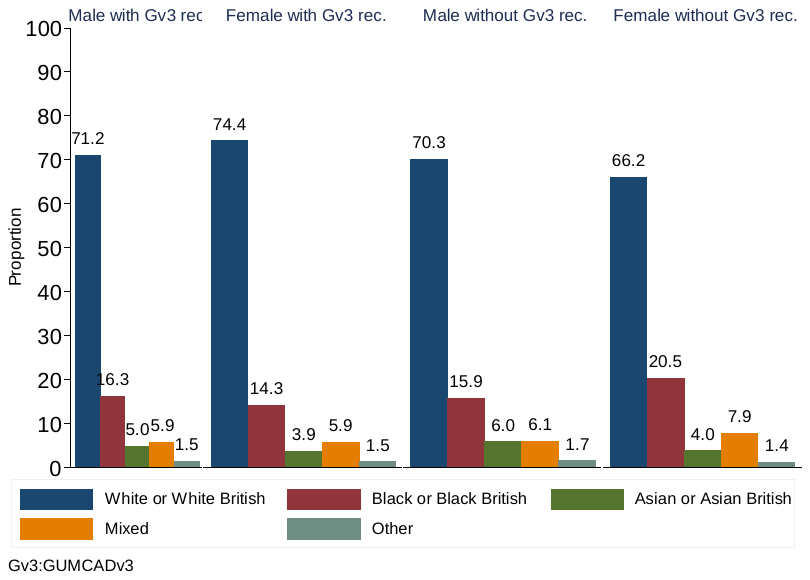

Key: GUMCADv3 – enhanced GUMCAD specification

The data collected in the pilot were based on a binary gender classification. The current version of GUMCAD collects data on gender identity (whether people are cisgender, transgender or gender-diverse).

Supplementary Material – Document 1. Topic guide for key informant interviews of staff participating in the first pilot of the enhanced GUMCAD specification

*GUMCADv3 is the enhancement of GUMCAD through the collection of behavioural and partner notification outcome variables. Your clinic recently participated in a pilot to assess the feasibility of collecting these additional items, and I would like to get more of your thoughts on this, on how successful the pilot was, and how we could possibly improve it.*

*Request permission to audio-record*:	**Y	N**

Date (dd/mm/yy):___________

Clinic:_____________________

Position: ___________________

i. In your opinion, how did the pilot go at your clinic? Was there any resistance from staff?
   - Staff engagement
   - Impact on routine work
   - Patient engagement
   - Completion of proforma
ii. Are the GUMCADv3 items usually collected during sexual history consultations? Were any questions awkward or difficult to ask?
iii. Which questions/domains do you think worked well, and why?
iv. Which questions/domains do you think did not work well, and why?
v. How did patients respond to being asked the questions:
   - On drug use
   - On risk behaviours (MSM online)
vi. What do you think would be the utility of these data?
    - Feasibility of long-term implementation
    - Assessment of risk behaviours
